# Cognitive Dysfunction in Non-CNS Metastatic Cancer: Comparing Brain Metastasis, Non-CNS Metastasis, and Healthy Controls

**DOI:** 10.1101/2022.11.10.22282138

**Authors:** Christopher Collette, Gabrielle Willhelm, Victor A. Del Bene, Stephen L. Aita, Dario Marotta, Terina Myers, Joseph Anderson, Meredith Gammon, Adam Gerstenecker, L. Burt Nabors, John Fiveash, Kristen L. Triebel

## Abstract

**Objective:** Cognitive impairment in primary and metastatic brain cancers has been well-documented. However, there is a lack of research comparing the cognitive profiles of people with non-central nervous system (CNS) metastatic cancer versus metastatic brain cancer.

**Methods:** This cross-sectional study consisted of 40 non-CNS metastasis, 61 brain metastasis, and 37 healthy control (HC) participants completing the same neuropsychological test battery.

**Results:** Both clinical groups had reduced processing speed, verbal learning/memory, and executive functioning. Non-CNS metastasis participants performed below HC participants on processing speed and executive functioning, while brain metastasis participants demonstrated lower performance across all neuropsychological tests. Semantic verbal fluency differentiated the two clinical groups (non-CNS metastasis>brain metastasis). Twenty-five percent of non-CNS metastasis participants and 57% of brain metastasis participants had ≥3 impaired scores (i.e., ≤5^th^ %ile).

**Conclusion:** One-quarter of non-CNS metastasis participants were cognitively impaired on at least three neuropsychological tests, and over half of brain metastasis participants demonstrated the same level of cognitive impairment. The elevated rate of cognitive dysfunction in the non-CNS metastasis participants is possibly attributable to systemic illness and treatment effects, while the cognitive deficits among brain metastasis participants may be associated with the more significant neurologic disease burden posed by brain metastases in conjunction with treatment effects.

## Introduction

The National Cancer Institute estimates that over 38% of all Americans will be diagnosed with cancer in their lifetimes. This percentage translates to over 1.7 million newly diagnosed cases per year (1). Roughly half will present with evidence of metastasis; of this population, 10% will present with brain metastasis (1-4). Where and when metastasis develops depends largely on the primary tumor location, genetic markers, and metabolic characteristics (3). Metastases are challenging to treat and are estimated to be responsible for 90% of cancer deaths (5). Brain metastases are associated with significant morbidity and mortality and an overall median survival time of just over seven months. However, some people with brain metastasis will live for years after diagnosis depending on the type of primary cancer (5).

Brain metastases are associated with significant cognitive impairments, affecting the quality of life (QoL), functional independence, and caregiver burden. In a randomized Phase III clinical trial of heterogeneous primary cancer patients with brain metastases, 90% of brain metastasis participants displayed cognitive impairment on at least one neurocognitive test, with most impairments observed in memory, executive functioning, and fine motor control (6). Similar rates of baseline cognitive impairment among people with brain metastasis have been demonstrated, with memory deficits present in over half of individuals (7-10). Additionally, those with brain metastasis have increased susceptibility to depression and anxiety (11), affecting cognition and QoL (12, 13).

Even among cancer patients without brain metastasis, cognitive impairment can be present before chemotherapy (14-17), occurring in up to 33% of people with breast cancer (17). A recent review of longitudinal research in people with breast cancer, pre- and post-chemotherapy, found impairments in memory, executive function, processing speed, and attention (18). High rates of cognitive impairment in pre-chemotherapy cancer participants suggest that chemotherapy is not the sole cause of cancer-related cognitive impairment (CRCI). Even rates of subjective and objective cognitive changes post-chemotherapy vary significantly in the literature from months to decades (19, 20). However, the literature on pre-chemotherapy cognitive impairment is scarce.

Literature is also sparse regarding CRCI studies in individuals with non-CNS metastatic cancer, which is vital for developing coping and treatment strategies and improving overall QoL. One such study (21) compared cognition and rates of impairment among localized colorectal cancer (n=281), metastatic colorectal cancer (n=66), and healthy controls (n=72) across ten clinical tests, and 47% of their patients with metastatic cancer had deficits in two or more domains compared to 15% of their healthy controls. Common domains with deficits included processing speed, attention, working memory, and verbal learning (21). This study uses neuropsychological tests to characterize the pattern and prevalence of cognitive impairment of non-CNS metastasis and brain metastasis. We hypothesized that both cancer groups would display cognitive impairment in multiple domains compared to healthy controls, with brain metastasis participants having a more severe cognitive impairment and at a higher frequency, due to the presence of brain tumor(s). We also expected both brain metastasis and non-CNS metastasis groups to show a similar profile of cognitive impairment as previously reported (7).

## Materials and Methods

### Participants

Newly diagnosed non-CNS metastasis (*n*=40) and brain metastasis (*n*=61) participants were recruited from the Department of Radiation Oncology and the Division of Hematology-Oncology between 2013 and 2018. A board-certified radiation oncologist made all diagnoses; eligible participants were 19 years or older. All brain metastasis participants had newly diagnosed first-time supratentorial lesions. Clinical participants with any history of one or more primary brain tumors, prior cranial radiation, leptomeningeal disease, neurological or psychiatric illness, substance abuse, or serious coexisting medical illness adversely affecting cognition were excluded from this study. Brain metastasis participants were assessed within one week of starting radiation therapy on the brain.

The HC group was comprised of healthy adult volunteers (*n*=37) recruited in a prior study (22). The HC group met the same eligibility requirements, except no history of cancer was permitted. They were recruited from the community through advertisements and were screened via structured telephone interviews to assess study appropriateness. None of the HC participants reported any cognitive symptoms.

The following treatments for brain metastases were used: surgical resection, single fraction radiosurgery with Gamma Knife or LINAC technology (15-24 Gy) for tumors ≤ 4 cm, hypofractionated focal radiation with LINAC for tumors > 3-4 cm (5-6 Gy x 5 fractions for 25-30 Gy total), and whole-brain radiation therapy (with LINAC; 30Gy in 10 fractions to 37.5 Gy in 15 fractions). Off-study guidelines for radiosurgical treatment followed the maximum tolerated doses outlined in RTOG 9005 (23). The majority of participants in the cancer groups did not have any surgical history—only seven brain metastasis (11%) and two non-CNS metastasis patients (5%) had prior resective surgery.

This study was approved by the University of Alabama at Birmingham Institutional Review Board (IRB 141023002) and was in accordance with the Declaration of Helsinki. All participants provided written informed consent.

### Measures

Commonly used neuropsychological tests were administered (see Table 1), including digit span (24), digit symbol subtest (24), verbal fluency (CFL/Animals) (25), Trail Making Test (TMT) Parts A and B (26), and the Hopkins Verbal Learning Test-Revised (HVLT-R) (27). The Karnofsky Performance Status (KPS) scale was used to measure functional status (28). The Hospital Anxiety and Depression Scale (29) and Beck Depression Inventory-II (30) were used to measure emotional distress in cancer and HC groups.

**Table 1.**
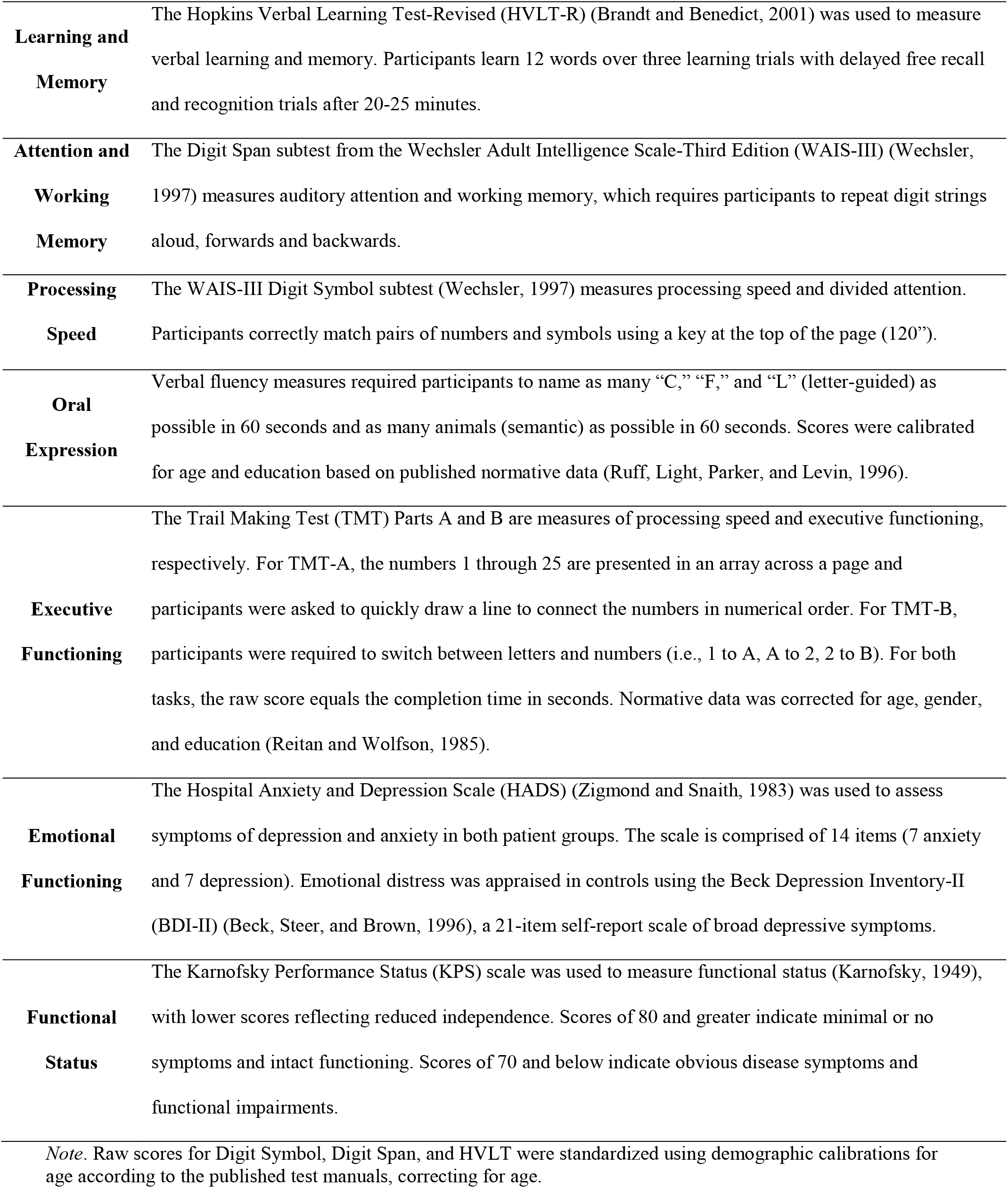
Description of Neurocognitive, Psychological, and Functional Measures

|  |  |
| --- | --- |
| **Learning and Memory** | The Hopkins Verbal Learning Test-Revised (HVLTR) (Brandt and Benedict, 2001) was used to measure verbal learning and memory. Participants learn 12 words over three learning trials with delayed free recall and recognition trials after 20-25 minutes. |
| **Attention and Working Memory** | The Digit Span subtest from the Wechsler Adult Intelligence Scale-Third Edition (WAIS-III) (Wechsler, 1997) measures auditory attention and working memory, which requires participants to repeat digit strings aloud, forwards and backwards. |
| **Processing Speed** | The WAIS-III Digit Symbol subtest (Wechsler, 1997) measures processing speed and divided attention. Participants correctly match pairs of numbers and symbols using a key at the top of the page (120”). |
| **Oral Expression** | Verbal fluency measures required participants to name as many “C,” “F,” and “L” (letter-guided) as possible in 60 seconds and as many animals (semantic) as possible in 60 seconds. Scores were calibrated for age and education based on published normative data (Ruff, Light, Parker, and Levin, 1996). |
| **Executive Functioning** | The Trail Making Test (TMT) Parts A and B are measures of processing speed and executive functioning, respectively. For TMT-A, the numbers 1 through 25 are presented in an array across a page and participants were asked to quickly draw a line to connect the numbers in numerical order. For TMT-B, participants were required to switch between letters and numbers (i.e., 1 to A, A to 2, 2 to B). For both tasks, the raw score equals the completion time in seconds. Normative data was corrected for age, gender, and education (Reitan and Wolfson, 1985). |
| **Emotional Functioning** | The Hospital Anxiety and Depression Scale (HADS) (Zigmond and Snaith, 1983) was used to assess symptoms of depression and anxiety in both patient groups. The scale is comprised of 14 items (7 anxiety and 7 depression). Emotional distress was appraised in controls using the Beck Depression Inventory-II (BDI-II) (Beck, Steer, and Brown, 1996), a 21-item self-report scale of broad depressive symptoms. |
| **Functional Status** | The Karnofsky Performance Status (KPS) scale was used to measure functional status (Karnofsky, 1949), with lower scores reflecting reduced independence. Scores of 80 and greater indicate minimal or no symptoms and intact functioning. Scores of 70 and below indicate obvious disease symptoms and functional impairments. |

*Note.* Raw scores for Digit Symbol, Digit Span, and HVLTR were standardized using demographic calibrations for age according to the published test manuals, correcting for age.

### Data Analysis

Differences in continuous (i.e., age and years of education) and categorical (i.e., gender and race) demographic variables were examined across groups using analysis of variance (ANOVA) and chi-squared test of independence, respectively. To describe the neurocognitive profiles, descriptive statistics (i.e., *M, SD*) were based on the standardized data across the 10 test scores. All standardized scores were transformed into *z*-scores via: 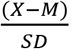

ANOVAs were used to examine neurocognitive test performances among the groups based on calibrated standardized scores. Eta squared (η^2^) was calculated for each ANOVA (small effect=.04, moderate effect=.25, and large effect=.64). For pairwise comparisons, Tukey’s Honestly Significant Difference (HSD) post hoc analysis was performed only on ANOVA models that were significant at the omnibus level. Finally, we examined cumulative impairment of test scores for the participant groups, defined as scores falling at or below the 5th%ile (i.e., *z*-score ≤ -1.64).

We then examined the base rate (i.e., prevalence) of cognitive impairment on seven unique tasks, producing ten total scores, looking at 0, ≥ 1, ≥ 2, and ≥ 3 impaired scores. Lastly, we examined proportional differences in rates of score impairment between the clinical groups using chi-squared tests of independence. The reference category was set to “0 impaired scores” for all the base rate and chi-squared analyses. Effect sizes were measured with the phi (ϕ) coefficient. Statistical analyses were conducted using IBM SPSS version 25.

## Results

### Sample Characteristics

Clinical groups did not significantly differ in age, gender, ethnicity, depression, or anxiety (see Table 2). The HC group (*M*=14.6, *SD*=1.8) had more education than the brain metastasis group (*M*=13.3, *SD*=2.6; on average, one more year of post-secondary education), but not the non-CNS metastasis group (*M*=13.8, *SD*=2.0). Since this difference in education was modest and not present between both clinical groups, this was not included as a covariate in our main findings since our primary interest was in the difference between non-CNS metastasis and brain metastasis groups. Further, analyses used normed scores calibrated for demographic factors (depending on the task). Nevertheless, we re-analyzed our data correcting for education and reported any discrepant results. Primary tumor locations and KPS data are in Table 3. In people with brain metastasis, 55.7% (34/61) had frontal lobe tumors, 21.3% (13/61) had temporal lobe tumors, and the remaining 23% (12/61) had mixed tumor locations. All three groups measured in the normal range of emotional distress.

**Table 2.** Participant Demographic Characteristics Stratified by Group

| Variable *M(SD)* | Controls(*n*=37) | Non-CNS<br>metastasis(*n*=40) | Brain<br>metastasis(*n*=61) | *p* |
| --- | --- | --- | --- | --- |
| Age | 56.8(12.1) | 59.7(12.3) | 59.8(11.6) | .436<sup>a</sup> |
| Education | 14.6(1.8) | 13.8(2.0) | 13.3(2.6) | .031<sup>a</sup> |
| HADS-D | -- | 4.3(3.0) | 5.4(4.0) | .17<sup>b</sup> |
| HADS-A | -- | 5.7(4.2) | 6.7(4.2) | .25<sup>b</sup> |
| BDI-II | 3.9(4.8) | -- | -- | -- |
| Gender *n*(%) |  |  |  |  |
| Male | 17(45.9) | 21(52.5) | 32(52.5) | .794<sup>c</sup> |
| Female | 20(54.1) | 19(47.5) | 29(47.5) |  |
| Race *n*(%) |  |  |  |  |
| Caucasian | 31(83.9) | 30(75.0) | 47(77.0) | .702<sup>c</sup> |
| African American | 6(16.2) | 10(25.0) | 13(21.3) |  |
| Other | -- | -- | 1(1.6) |  |

**Abbreviations:** HADS-D and HADS-A= The Hospital Anxiety and Depression Scale-depression and anxiety subscale, respectively. <sup>a</sup>*p*-values represent ANOVAs for age and education. <sup>b</sup>*p*-values represent independent *t*-tests for HADS-D and HADS-A. <sup>c</sup>*p*-values represent chi-square tests for gender and race.

**Table 3.** Functional Status and Primary Tumor Location

| Variable | Non-CNS | Brain metastasis(*n*=61) |
| --- | --- | --- |
|  | metastasis(*n*=40) |  |
| Karnofsky Score | *n*(%) |  |
| 100-80(Able to Work) | 28(70) | 46(75.4) |
| 70-50(Unable to Work) | 12(30) | 14(23.0) |
| 40-0(Functionally Disabled) | -- | 1(1.6) |
| Primary Tumor Location | *n*(%) |  |
| Lung<sup>1</sup> | 9(22.5) | 32(52.5) |
| Breast | 6(15.0) | 10(16.4) |
| Melanoma | 3(7.5) | 4(6.6) |
| Gynecological<sup>2</sup> | 3(7.5) | 5(8.2) |
| Renal Cell | 1(2.5) | 2(3.3) |
| Gastrointestinal<sup>3</sup> | 10(25.0) | 4(6.6) |
| Prostate | 5(12.5) | 1(1.6) |
| Testicular | -- | 1(1.6) |
| Other | 3(7.5) | 2(3.3) |

<sup>1</sup>Non-CNS: 9 non-small cell; Brain: 29 non-small cell, 3 small cell.
<sup>2</sup>Non-CNS: 1 gynecological, 1 ovarian, 1 cervical; Brain: 2 gynecological, 2 ovarian, 1 cervical.
<sup>3</sup>Non-CNS: 2 pancreatic, 1 gallbladder, 1 liver, 1 rectum, 5 colon; Brain: 2 pancreatic, 1 gastric, 1 colon.

Overall, 77.5% of non-CNS metastasis participants received chemotherapy in the past compared to 37.7% of brain metastasis participants. Additionally, 45.0% of non-CNS metastasis participants were receiving chemotherapy at the time of the study compared to 13.1% of brain metastasis participants. Among cancer participants, lifetime history of chemotherapy (past and/or present) was not significantly associated with cognitive performance, while to our surprise, current chemotherapy treatment was weakly positively associated with higher cognition across tasks (Pearson product-moment correlation coefficients values ranging from .13 to .35). Surgical resection was more common among brain metastasis participants (11.5%) than non-CNS metastasis participants (5.0%). Comparable percentages of non-CNS metastasis (70.0%) and brain metastasis (65.6%) participants started radiation therapy within a week of the study. All brain metastasis participants had supratentorial lesions. Of the brain metastasis group, 41.0% had 1 metastasis, 32.8% had 2-3 metastases, and 26.2% had > 3 metastases. Functional ability was comparable between non-CNS and brain metastasis groups (median KPS score=80), and clinical participants were functionally intact on average. Eighteen HC (48.6%) were missing HVLT-R Delayed Recall and Retention data, but all these participants had at least HVLT-R Recognition Discriminability Index scores. The data were considered missing at random as there were no systematic statistical differences between controls with and without missing data. Further, this missing data did not interfere with our primary goal of examining the prevalence of score impairment and differences between the cancer groups. Nine brain metastasis participants (14.8%) had incomplete neuropsychological test data, though the majority (66.7%) were only missing one of ten task scores. Missing data in the ANOVA models were handled via pairwise deletion.

### Cognitive Performance

Examination of the neuropsychological profile for the clinical groups showed normatively low average-to-average performances across all tasks at a group level (see Table 4). However, non-CNS metastasis participants performed better than brain metastasis participants across all tasks. Both clinical groups showed preserved basic attention and working memory ability on the Digit Span task and information processing speed on TMT-A, with comparable rates of impairment seen in a subgroup of individuals. Conversely, the clinical groups displayed lower mean performance on Digit Symbol Coding, a more complex task of rote visual learning and speed of mentation, with high rates of score impairment (non-CNS=22.5%; brain=33.3%). Though the clinical groups generally had average mean performance and low rates of score impairment on Semantic Fluency (non-CNS=5.0%; brain=12.7%), performance on Letter-Guided Fluency was lower, with the brain metastasis group showing a markedly higher base rate of score impairment (non-CNS=12.5%; brain=34.9%). The clinical groups showed normatively lower performance on verbal learning (HVLT-R Total Recall: non-CNS=20.0%, brain=36.5%) and delayed memory (HVLT-R Delayed: non-CNS=15.0%, brain=36.5%), with brain metastasis participants having the highest prevalence of score impairment. Both clinical groups showed non-trivial base rates of score impairment on HVLT-R Retention (non-CNS=10.0%; brain=15.9%) and HVLT-R Recognition (non-CNS=12.5%; brain=15.9%). Regarding executive functioning, the clinical groups displayed relatively worse normative performance on TMT-B, with the brain metastasis group having a higher base rate of impaired scores (non-CNS=12.5%; brain=23.8%).

**Table 4.** Neurocognitive Performance as a Function of Group and Domain

| Measures by Domain | Standard-score (z): *M(SD)* | | | %Impaired | | | *F(df)*<sup>b</sup> | *p*($\eta^2$) | Post-hoc<sup>b,c</sup> |
| --- | --- | --- | --- | --- | --- | --- | --- | --- | --- |
|  | Controls | Non-CNS<br>metastasis | Brain<br>metastasis | Controls<sup>a</sup> | Non-CNS<br>metastasis | Brain<br>metastasis |  |  |  |
| **Attention** |  |  |  |  |  |  |  |  |  |
| Digit Span<sup>d</sup> | 0.17(0.76) | -0.04(0.88) | -0.23(0.72) | 0.0 | 5.0 | 4.8 | 3.1(2,134) | .048(.04) | C>Brain |
| **Processing Speed** |  |  |  |  |  |  |  |  |  |
| Digit Symbol Coding | 0.49(1.03) | -0.55(1.09) | -0.84(1.03) | 0.0 | 22.5 | 33.3 | 20.8(2,130) | <.001(.22) | C>N-CNS,Brain |
| TMT-A(Time) | 0.44(1.06) | 0.31(1.01) | -0.18(1.18) | 2.7 | 2.5 | 7.9 | 4.3(2,132) | .016(.06) | C<Brain |
| **Expressive Language** |  |  |  |  |  |  |  |  |  |
| Letter-Guided Fluency | -0.06(1.11) | -0.44(1.02) | -0.89(1.14) | 13.5 | 12.5 | 34.9 | 6.6(2,133) | .002(.09) | C>N-CNS,Brain |
| Semantic Fluency<sup>e</sup> | -0.04(0.87) | 0.23(1.08) | -0.35(1.25) | 0.0 | 5.0 | 12.7 | 3.3(2,133) | .038(.05) | N-CNS>Brain |
| **Memory** |  |  |  |  |  |  |  |  |  |
| HVLT-R Total Recall | -0.22(0.87) | -0.45(1.05) | -0.92(1.19) | 2.7 | 20.0 | 36.5 | 5.4(2,135) | .005(.07) | C>Brain |
| HVLT-R Delayed | -0.42(0.79) | -0.37(1.16) | -0.97(1.18) | 0.0 | 15.0 | 36.5 | 6.5(2,117) | .002(.10) | C>Brain |
| HVLT-R Retention(%) | 0.07(0.89) | 0.02(1.21) | -0.46(1.16) | 2.7 | 10.0 | 15.9 | 2.8(2,117) | .063(.04) | -- |
| HVLT-R Recognition | 0.27(0.70) | -0.09(1.01) | -0.35(1.04) | 2.7 | 12.5 | 15.9 | 4.9(2,135) | .009(.07) | C>Brain |
| **Executive Functioning** |  |  |  |  |  |  |  |  |  |
| TMT-B(Time) | 0.22(1.19) | -0.40(1.14) | -0.78(1.16) | 8.1 | 12.5 | 23.8 | 18.6(2,128) | <.001(.13) | C<Brain,N-CNS |

**Abbreviations:** TMT=Trail Making Test; HVLT-R=Hopkins Verbal Learning Test-Revised; C=Control Group; N-CNS=Non-CNS Metastasis Group;
Brain=Brain Metastasis Group. <sup>a</sup>18 Controls missing complete HVLT-R data; rates of impairment computed based on available data. <sup>b</sup>Analyses based
on standardized scores. <sup>c</sup>Post-hoc analyses reflect pairwise comparisons using Tukey's HSD for statistically significant ( $\alpha=.05$ ) comparisons. <sup>d</sup>Not
significant after correcting for education ( $p=.123$ , $\eta_p^2=.03$ ). <sup>e</sup>Not significant after correcting for education ( $p=.054$ , $\eta_p^2=.04$ ).

Next, test performances among the control and clinical groups were compared. Significant group differences in cognitive performance emerged for the Digit Span (*p*=.048, η^2^=.04), Digit Symbol Coding (*p*<.001, η^2^=.22), TMT-A (*p*=.016, η^2^=.06) and TMT-B (*p*<.001, η^2^=.13), Letter-Guided Fluency (*p*<.002, η^2^=.12), Semantic Fluency (*p*=.038, η^2^=.05), HVLT-R Total Recall (*p*=.005, η^2^=.07), HVLT-R Delayed Recall (*p*=.002, η^2^=.10), and HVLT-R Recognition (*p*=.009, η^2^=.07) tasks. No significant group differences in HVLT-R Retention were observed. Post-hoc analyses showed that the brain metastasis group performed significantly lower than controls on Digit Span, Digit Symbol Coding, TMT-A and TMT-B, Letter-Guided Fluency, and the HVLT-R Total and Delayed Recall. Non-CNS metastasis participants performed lower than controls on Digit Symbol Coding, Letter-Guided Fluency, and TMT-B. Non-CNS metastasis participants had greater Semantic Fluency performance than the brain metastasis group. All ANOVAs were re-run with education added as a covariate, and results were identical except Digit Span (*p*=.123, η _p_^2^=.03) and Semantic Fluency (*p*=.054, η_p_^2^=.04).

Within the entire healthy control sample, 30 (81%) had no impaired scores, only four participants (10.8%) had one impaired score, and one person (2.7%) had two impaired scores. Lastly, the base rate of cumulative score impairment was compared between clinical groups (Table 5). Relative to non-CNS metastasis participants, more brain metastasis participants had ≥ 1 impaired scores across tests (70.5% vs. 47.5%; ϕ=.23). Stated differently, a little over half of the non-CNS metastasis (52.5%) and only around one-quarter of brain metastasis (29.5%) participants had no impaired scores. In addition, a significantly greater proportion of brain metastasis vs. non-CNS metastasis participants had ≥ 2 (65.4% vs. 38.2%; ϕ=.27) as well as ≥ 3 (57.1% vs. 25.0%; ϕ =.32) impaired test scores. As expected, the HC group had the lowest impairment rates across virtually all test scores, and the majority (81.1%) had no impaired scores.

**Table 5.** Frequency of Total Number of Impaired Cognitive Measures by Participant and Group

| # of Impaired<br>Tests<sup>†a</sup> | Controls<sup>b</sup> | Non-CNS | Brain | $\chi^2(\phi)^d$ | *p* |
| --- | --- | --- | --- | --- | --- |
|  |  | metastasis | metastasis<sup>c</sup> |  |  |
|  |  | *n*(%) |  |  |  |
| 0 | 30(81.1) | 21(52.5) | 18(29.5) | -- | -- |
| ≥ 1 | 7(18.9) | 19(47.5) | 43(70.5) | 5.39(.23) | .020 |
| ≥ 2 | 3(8.1) | 13(38.2) | 34(65.4) | 6.11(.29) | .013 |
| ≥ 3 | 2(5.4) | 7(25.0) | 24(57.1) | 7.04(.32) | .008 |

<sup>†</sup>Impaired at ≤ 5th percentile (*z*-score ≤ -1.64); based on a battery of 10 test scores unless otherwise specified.
<sup>a</sup>'0 impaired tests' was set as the comparison category for chi-square analyses.
<sup>b</sup>9 brain metastasis participants did not have complete data; rates of impairment were based on available data.
<sup>c</sup>18 HC had partial HVLT-R data; rates of impairment were based on available data.
<sup>d</sup>Chi-square analyses based on Non-CNS and Brain metastasis groups.

## Discussion

This current study found cognitive impairment in participants with non-CNS metastatic cancer and compared their cognitive profiles to brain metastasis and healthy control groups. As expected, brain metastasis participants had higher rates of cognitive impairment across all domains. Rates of impairment among non-CNS metastasis participants were attenuated and generally limited to the processing speed and executive functioning domains but were present in a quarter or more of individuals, depending on the number of impaired scores. While at a group level, the mean scores for the non-CNS and brain metastasis groups were mainly in the low average-to-average ranges, significant, non-trivial portions of both groups demonstrated varying rates of cognitive impairment. Twenty-five percent of non-CNS metastasis and 57% of brain metastasis participants had impaired scores on three or more tests. This rate of cognitive impairment is consistent with prior findings of pre-treatment cancer affecting cognition in cancer participants with and without brain metastasis (5-7, 10, 12, 15, 18, 31-33).

Broadly, this study investigated cognitive impairment in people with non-CNS metastatic cancer. Cognitive impairment in cancer is likely multifactorial, related to factors of systemic illness and treatment effects (16-18, 33), though we did not directly examine these factors. Most of our non-CNS metastasis group had previously received chemotherapy, and 45% were receiving chemotherapy at the time of this study. While chemotherapy may contribute to the CRCI in our non-CNS metastasis group, our study found the most considerable rates of cognitive impairment in the brain metastasis group, despite lower rates (13%) of chemotherapy. Furthermore, past and/or present chemotherapy history was only modestly associated with cognitive data. In addition, all patients in our study were assessed within one week of starting radiation treatment. While radiation treatment can affect cognition (6, 8, 34), this is typically observed in 30% or more of patients after four months of radiation treatment (partial or whole brain), which is well beyond our one-week time window. Moreover, for chemotherapy, the literature does not strongly indicate a significant decline in cognitive functioning within this short time interval (19). In the brain metastasis group, the higher rate of cognitive impairment is not surprising due to the presence of metastasis in the brain.

Both metastasis groups had elevated rates of deficits in processing speed and executive functioning. The brain metastasis group also demonstrated deficits in attention and memory, consistent with prior studies (9, 10, 15, 16, 18). Relative to our healthy control group, our two clinical groups had higher impairment rates on all tests except for letter-guided verbal fluency, with no differences in impairment rates observed between the healthy control and non-CNS metastasis participants (13.5% and 12.5%, respectively). In contrast, 33.3% of the brain metastasis group met the previously established criteria for an impaired letter-guided verbal fluency score. When comparing our observed impairment rates to a study of colorectal cancer with metastasis conducted by Vardy and colleagues (21), our healthy control participants had notably lower rates of impairment across overlapping tests (e.g., Digit Span, Symbol Digit, TMT-A & B, and HVLT-R). While our HC sample impairment ranged from 0% to 8.1%, their HC sample impairment rates ranged from 14% to 19%. Their metastatic cancer group’s impaired scores ranged from 15% to 47%, while our non-CNS metastasis group ranged from 5% to 22.5%, and our brain metastasis group ranged from 4.8% to 36.5%. This discrepancy is likely due to their more liberal cutoff score (T-Score = 40 or 16^th^ %ile), while we used a more conservative cutoff at the 5^th^ %ile. A more liberal cutoff will inflate impairment rates. To demonstrate this point, in a study examining how antiangiogenic treatment affects individuals with metastatic renal cancer, pre-treatment rates of cognitive impairment were highly similar to the rates observed in our sample when the same conservative cutoff was applied (35).

The impairments observed in both our clinical groups are possibly driven by frontostriatal dysfunction (7), with frontostriatal projections related to attention, processing speed, working memory, and executive functioning (36-38). While cancer-related treatment does affect cognition, there is evidence to suggest that pre-chemotherapy, women who are diagnosed with breast cancer (stages I-III) have deficits in attention and working memory, along with an atypical pattern of increased hemodynamic response in anterior cingulate and inferior frontal gyrus as measured with functional MRI. Moreover, those who demonstrated greater task difficulty also had greater BOLD activation across more distributed brain regions (39), suggesting inefficient neural processing. Several other studies have shown that pre-chemotherapy individuals with cancer have aberrant patterns of brain activity (40-43), reduced white matter integrity (42), and reduced cognitive functioning (14, 15, 42). However, one study found the typically observed increased hemodynamic pattern in several brain regions (inferior frontal gyrus, insula, thalamus, midbrain) but no evidence of cognitive changes on a visuospatial n-back test designed to target working memory (43). Some, but not all, of these cognitive and functional imaging changes appear to be partially related to depression, anxiety, and fatigue in early-stage breast cancer (41). Thus, the presence of cancer appears to have an indirect effect on brain function that is not explained by these factors.

Individuals with cancer commonly experience depression (11), and as described above, it could be related to brain function in people with cancer (41). To our surprise, our brain and non-CNS metastasis groups did not report different levels of depression on the HADS. However, we could not statistically test for group differences with HC participants because they were administered the BDI-II, but all three groups measured in the normal range on average. Therefore, mood symptoms affecting cognitive performance in our sample are unlikely.

While at the group level, our cancer patients did not demonstrate frankly impaired scores (normatively), there are clinical implications stemming from the high base rate of cumulative score impairment observed. An important consideration for patients, caregivers, and treatment teams is that functional capacity and medical decision-making are vulnerable to the combined effects of cancer and age-related cognitive impairments (22, 44). In a recently published study, 60% of individuals with brain metastasis and 54% of non-CNS metastasis participants showed reduced capacity to consent to research (45). Along these lines, people with brain and non-CNS metastasis who demonstrate cognitive deficits are at a higher risk of not comprehending medical treatments (44). Hence, assessing cognition in cancer participants is crucial, especially in advanced metastatic cancer. Understanding the cognitive sequelae associated with brain and non-CNS metastatic cancer has substantial implications for the individual, their treatment team, and their caregiver network. Our findings can help inform expectations and focus on remediation and compensatory strategies. Education should be provided with the individual’s cognitive capabilities in mind. When possible, caregivers should be present since the observed cognitive impairments may interfere with their understanding and recall of medical information. Additionally, the treating team should regularly inquire about caregiver burden since this increases in the context of cognitive decline (46), which is present in a large percentage of both brain and non-CNS metastatic cancer groups.

Several factors limit the current research. Participants were recruited from a single academic medical center, and our sample was relatively small, which may have reduced the generalizability, although our clinical samples are well characterized. In addition, our sample is comprised of heterogeneous solid-based primary tumors in a cross-sectional design, which precludes causal inference. It is unclear if metastatic cognitive profiles differ as a function of primary cancer types. However, the prior literature shows that cancer, brain metastases, and treatment are all pathways to cognitive decline. Assessing cognition with different primary tumor types may be informative as to how sociodemographic factors relate to cognition in the brain and non-CNS metastasis participants. While our clinical groups did not report elevated rates of depression, other factors not measured, such as fatigue, could have possibly influenced cognition, functional decline, and disability and should be further investigated. Although a sizable proportion of the HC group had incomplete HVLT-R data, this did not detract from our primary research aim. Furthermore, this did not occur systemically, and no significant differences in scores were observed between the three groups.

## Conclusion

In summary, the presence of metastatic cancer, both with and without direct involvement in the central nervous system, is associated with a higher prevalence of cognitive dysfunction. This study contributes to the relatively small number of investigations looking at cognition in non-CNS metastatic cancer. In individuals with metastatic brain cancer, all cognitive domains showed diminishment. Furthermore, these individuals had higher rates of cognitive impairment, with deficits in processing speed and executive functioning observed in over a quarter of individuals. While the non-CNS metastatic cancer group had lower rates of impairment than the metastatic brain group, 25% still demonstrated cognitive impairment on three or more tests, which has important clinical implications on treatment planning, consenting for treatment and research, quality of life, and caregiver burden. Our findings can help inform individuals with metastasis, their treatment team, and caregivers.

## Data Availability

All data produced in the present study are available upon reasonable request to the authors

## Notes

**Conflict of Interest:** None declared

**Funding:** This work was supported by the American Cancer Society [MRSG-14-204-01 to KT]; the National Institutes of Health/National Center for Advanced Translational Sciences [KL2 TR000166 to KT]; the National Cancer Institute [5R25CA076023]; and the University of Alabama at Birmingham Department of Neurology.

### Competing Interest Statement

The authors have declared no competing interest.

### Funding Statement

This work was supported by the American Cancer Society [MRSG-14-204-01 to KT]; the National Institutes of Health/National Center for Advanced Translational Sciences [KL2 TR000166 to KT]; the National Cancer Institute [5R25CA076023]; and the University of Alabama at Birmingham Department of Neurology.

### Author Declarations

This study was approved by the University of Alabama at Birmingham Institutional Review Board (IRB 141023002)

